# A *KLHL40* 3’ UTR splice-altering variant causes milder NEM8, an under-appreciated disease mechanism

**DOI:** 10.1101/2022.08.08.22278402

**Authors:** Lein N.H. Dofash, Gavin Monahan, Emilia Servián-Morilla, Eloy Rivas, Fathimath Faiz, Patricia Sullivan, Emily Oates, Joshua Clayton, Rhonda L. Taylor, Mark R. Davis, Traude Beilharz, Nigel G. Laing, Macarena Cabrera-Serrano, Gianina Ravenscroft

## Abstract

Nemaline myopathy 8 (NEM8) is typically a severe autosomal recessive disorder associated with variants in the kelch-like family member 40 gene (*KLHL40*). Common features include fetal akinesia, fractures, contractures, dysphagia, respiratory failure, and neonatal death. Here, we describe a man in his 20s with relatively mild NEM8. He presented with hypotonia and bilateral femur fractures at birth, later developing bilateral Achilles’ contractures, scoliosis, and elbow and knee contractures. He had walking difficulties throughout childhood and became wheelchair bound during adolescence after prolonged immobilisation. Muscle MRI during adolescence indicated prominent fat replacement in his pelvic girdle, posterior compartments of thighs, and vastus intermedius. Muscle biopsy revealed nemaline bodies and intranuclear rods. RNA sequencing and western blotting of patient skeletal muscle indicated significant reduction in *KLHL40* mRNA and protein respectively. Using gene panel screening, exome sequencing and RNA sequencing, we identified compound heterozygous variants in *KLHL40*; a truncating 10.9 kb deletion *in trans* with a likely pathogenic variant (c.*152G>T) in the 3’ untranslated region (UTR). Computational tools SpliceAI and Introme predicted the c.*152G>T variant created a cryptic donor splice site. RNA-seq and *in vitro* analyses indicated that the c.*152G>T variant induces multiple de novo splicing events that likely provoke nonsense mediated decay of *KLHL40* mRNA explaining the loss of mRNA expression and protein abundance in the patient. Analysis of 3’ UTR variants in ClinVar suggests SNPs that introduce aberrant 3’ UTR splicing may be underrecognised in Mendelian disease. We encourage consideration of this mechanism during variant curation.

## Introduction

Nemaline myopathies (NEM) are a clinically and genetically heterogenous group of congenital myopathies characterised by the presence of nemaline bodies within muscle fibres(1). Clinical symptoms include muscular hypotonia and weakness, often accompanied by respiratory insufficiency(2). The age of onset, disease progression and severity are variable and largely depend on the underlying genetic cause(2). To date, 13 genes have been associated with NEM(3) and are routinely screened in diagnostic laboratories(4). Generally in diagnostic laboratories, analysis of known disease genes is restricted to rare coding and structural variants, while variants in regulatory regions are often uncaptured or overlooked(5).

One common severe subtype of NEM is nemaline myopathy 8 (NEM8; OMIM# 615348), an autosomal recessive disorder associated with biallelic variants in the kelch-like family member 40 gene (*KLHL40*)(6). Almost all patients with NEM8 present with severe disease(6, 7). Characteristic features include fetal akinesia or hypokinesia, fractures, contractures, facial involvement, dysphagia, respiratory failure, and neonatal death (average age at death in a NEM8 cohort was 5 months of age)(6). To date, only two patients have been reported with relatively mild NEM8, one of whom responded beneficially to acetylcholinesterase inhibitors(8, 9).

KLHL40 is a member of the kelch-repeat-containing protein superfamily and is involved in a range of interactions that regulate skeletal muscle myogenesis and promote skeletal muscle maintenance(10, 11). KLHL40 is reported to bind the E2F1-DP1 complex to regulate myogenesis(11). In addition, KLHL40 binds NEB and LMOD3 to stabilise the thin filament(10). To date, pathogenic variants that change the *KLHL40* open reading frame have been implicated in NEM8(6, 7). These include protein truncating variants such as frameshift, essential splice site, and nonsense variants(6). Pathogenic missense variants have also been reported and are predicted to destabilise KLHL40 by disrupting intramolecular interactions(6). KLHL40 deficiency consequently destabilises the thin filament and causes nemaline myopathy(10). NEM8 patients typically show reduced or absent KLHL40 in skeletal muscle by western blotting(6-8).

In this study, we describe a Spanish patient with mild NEM8 who had diminished *KLHL40* transcript and protein abundance. He harbours a multi-exon KLHL40 deletion on one allele and a splicing-altering SNP in the 3’ UTR of *KLHL40* on the other allele. The 3’ UTR SNP creates a cryptic donor splice site 152 bp downstream of the termination codon, likely inducing nonsense mediated decay. Our analysis of ClinVar Class 3-5 variants, suggests this 3’ UTR mechanism may be underrecognised in Mendelian disease.

## Results

### Clinical features and pathology

The patient is a male born to non-consanguineous parents, with no relevant family history of disease. He was born hypotonic with bilateral femur fractures. He had delayed walking and was never able to jump or run. He lost independent ambulation during adolescence, after prolonged immobilization after Achilles tendon lengthening. From childhood he had scoliosis, a myopathic face and ogival palate, and weakness of bilateral orbicular oculi muscles. He was last examined during early adulthood, being able to stand with bilateral support. At last examination, he had weakness of proximal and distal muscles of upper and lower limbs and showed obvious atrophy of intrinsic hand muscles. Deep tendon reflexes were absent, with bilateral Achilles tendon contractures as well as elbow contractures and cervical and dorsal rigid spine.

A muscle MRI showed widespread fat replacement of pelvic girdle muscles, thighs, and lower legs (Figure 1A-C). At the thigh level, the fat replacement was particularly severe involving posterior compartments as well as *rectus anterioris*.

**Figure 1.**
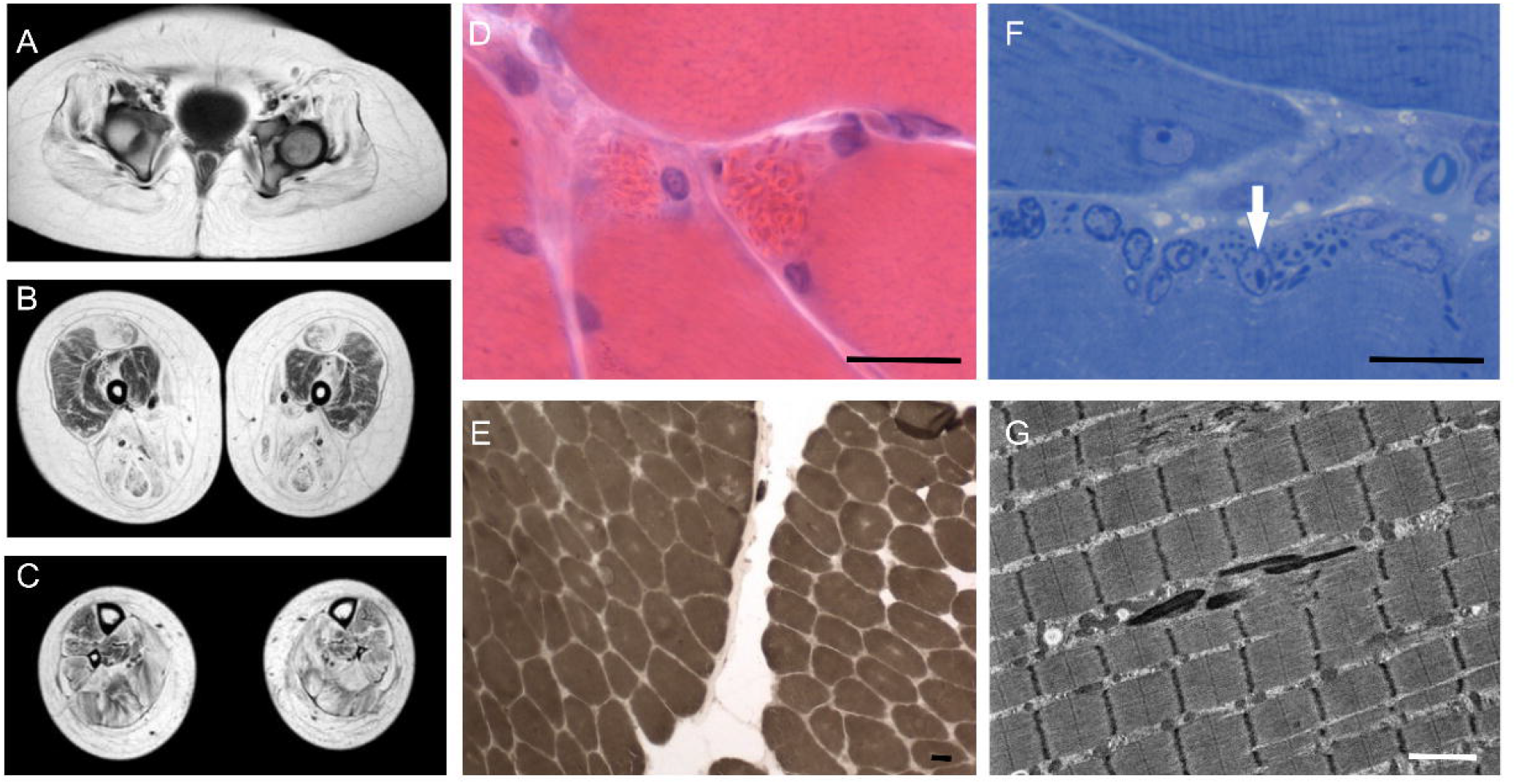
Muscle pathology in the patient with a milder form of NEM8. (A,B,C) Magnetic resonance imaging shows extensive fatty replacement of muscles of pelvic girdle (A), thighs (B), with most prominent involvement of posterior compartment (B) and legs (C). Muscle biopsy sections: H&E stain showing clusters of nemaline bodies (D), ATPase 4.3 stain demonstrating all myofibres in the sample are type 1 (E), Semithin slide stained with toluidine blue showing an intranuclear rod (F, arrow), Electron micrograph displaying electron-dense rod-shaped structures that correspond to nemaline bodies (G). Scale bar: 20 µm (D-F); 2 µm (G).

Light microscopy examination revealed moderate and patchy fat replacement, without relevant fibrosis, with mild variability in fibre size. Necrotic or regenerating fibres were not seen. Numerous eosinophilic birefringent structures, rod or round shaped, were seen forming clusters in the centre of the sarcoplasm or subsarcolemmal regions (Figure 1D). There were internal nuclei. Some intranuclear rods were identified on the semithin sections (Figure 1F). Gomori trichrome confirmed the presence of abundant rods with irregular distribution in the sarcoplasm (not shown). Histoenzymatic techniques showed an irregular intermyofibrillar pattern, with presence of pseudo-lobulated fibres and some core-like areas devoid of oxidative activity harbouring the clusters of rods. ATPase stains showed 100% of the fibres were type 1 (Figure 1E).

Electron microscopy confirmed the presence of numerous electron-dense rod-shaped structures corresponding to nemaline rods in most muscle fibres (Figure 1G). Rods were irregularly distributed throughout the fibre, clustered centrally and formed subsarcolemmal clusters in the periphery. They often ran parallel to the long axis of the sarcomere and showed continuity with Z-disks.

### Genomic, transcriptomic and protein investigations

Gene panel sequencing and whole exome sequencing (WES) identified a heterozygous 10.9 kb deletion in *KLHL40* (hg19; chr3:42729537-42740458) in the patient (Figure 2A). This deletion spans exons 2-6 of the six exons of *KLHL40* (ENSG00000157119) and results in complete loss of a functional allele. The deletion was absent in gnomAD (v2.1) and was only present in this patient in the 10,000 alleles in the Broad structural variant callset(12).

**Figure 2.**
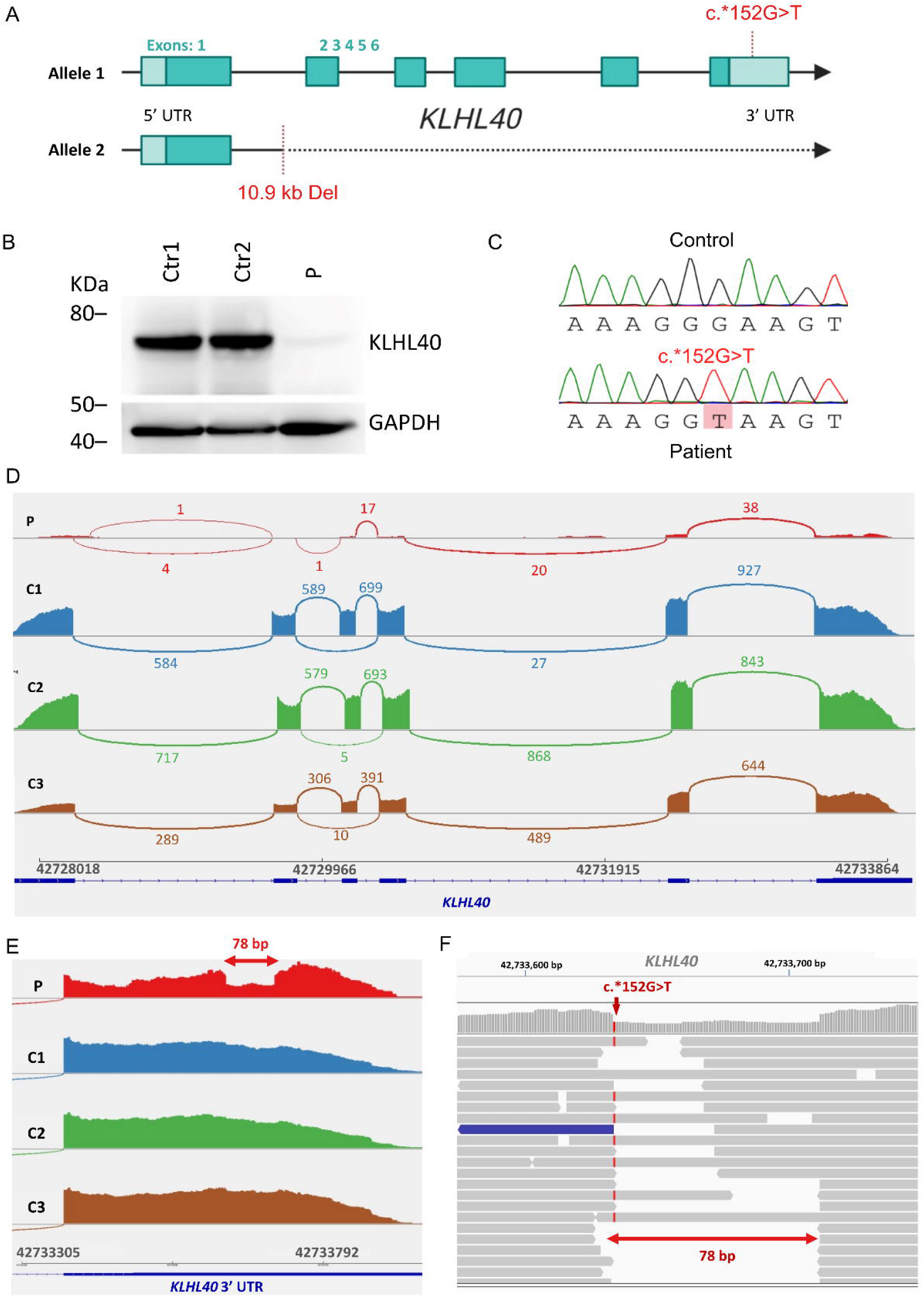
Alterations of *KLHL40* at the DNA, mRNA, and protein level in the patient. (A) Schematic representation of the *KLHL40* gene with the 5’ and 3’ UTRs (light blue), 6 coding exons (cyan) and the compound heterozygous variants in the patient. Figure designed with BioRender.com. (B) Western blotting for KLHL40 (top panel) from patient (P) and control (Ctrl 1-2) muscle biopsies. Western blotting for GAPDH (bottom panel) as a loading control. (C) Sanger confirmation of the c.*152G>T variant in Patient (bottom) compared to an unaffected control (top), the variant appears homozygous due to deletion on the second allele. (D-F) RNA-seq data of muscle RNA from the patient and controls C1-C3 aligning to *KLHL40*. (D) Sashimi plots for patient and controls. (E) Close up of Sashimi plots at the 3’ UTR region of *KLHL40*. (F) Patient RNA-seq reads spanning the 3’ UTR region with the c.*152G>T variant. Sequences viewed using the Integrative Genomics Viewer (IGV).

Western blotting indicated very low abundance of KLHL40 in patient skeletal muscle compared to healthy controls (Figure 2B). Although deficiency of KLHL40 is sufficient for diagnosis of NEM8, a second variant *in trans* would be expected given the recessive nature of KLHL40-related nemaline myopathy(6). Following re-analysis of WES data to include rare non-coding variants, a single nucleotide variant (c.*152G>T) was identified in the 3’ UTR of *KLHL40 in trans* with the deletion (Figure 2A). The variant appeared homozygous given the complete absence of the 3’ UTR on the other allele harbouring the exon 2-6 deletion. The c.*152G>T variant was present in gnomAD (v3.1.2) at a frequency of 3.94 ×10^−5^ (6/152,204 alleles; no homozygotes). The c.*152G>T variant was confirmed by bidirectional Sanger sequencing (Figure 2C). Parental DNAs are not available.

The SpliceAI tool(13) predicted that the c.*152G>T variant creates a weak donor splice site which results in a 255 bp cryptic intron in the 3’ UTR of *KLHL40* (positions c.*151 to c.*405). The donor gain prediction score (0.17) was below the Δ score threshold (0.2) suggested for SpliceAI (13). Nevertheless, this prediction was supported by the Introme tool(14) which had a score above the threshold (Introme score 0.76, threshold 0.54).

Analysis of patient skeletal muscle RNA-seq data showed significantly lower levels of *KLHL40* transcripts compared to non-nemaline myopathy patient skeletal muscle (control, Figure 2D). Patient RNA-seq coverage ranged between 1-38 reads (Figure 2D). Control RNA-seq coverage ranged between 5-927 reads (Figure 2D). The low coverage of *KLHL40* appeared specific in the patient sample, as IGV analyses indicated comparable coverage of 10,670 other genes in the NEM8 patient compared to controls. Normal splicing of *KLHL40* exons 1-6 was observed at low frequency (Figure 2D). Splicing of the predicted 255 bp cryptic intron was not detected by RNA-seq, although a distinct drop in coverage over 78 bp neighbouring the 3’ UTR variant site was observed (Figure 2E,F) suggesting the formation of a 78 bp cryptic intron which would result in a secondary variant in the 3’ UTR of *KLHL40*; c.*151_*228del. MINTIE(15) alignment of the RNA-seq reads confirmed the 78 bp deletion (hg19; chr3:42733635-42733714) and indicated the presence of a new exon junction with a variant allele frequency of 0.94. This implies that 94% of the reads contained the cryptic 78 bp deletion. Some RNA-seq reads also suggested the formation of other cryptic deletions, however these were not detected by MINTIE alignment.

### *In vitro KLHL40* expression analyses

To functionally demonstrate the effect of the c.*152G>T variant on *KLHL40* expression and abundance, we transiently expressed *KLHL40* with either the wild-type (WT) or mutant (c.*152G>T) 3’ UTR in HEK293FT cells. We also generated a 78bp deletion mutant (c.*151_*228del) to mimic the cryptic intron identified by RNA-seq.

Quantitative PCR analysis indicated significantly lower expression of the *KLHL40*^c.*152G>T^ construct (1.7 ± 0.1) relative to *KLHL40*^WT^ (2.4 ± 0.1) and *KLHL40*^c.*151_*228del^ (2.5 ± 0.1), confirming the association of the c.*152G>T variant with reduced *KLHL40* transcript levels (Figure 3A). The difference in expression between the *KLHL40*^WT^ and *KLHL40*^c.*152G>T^ constructs was statistically significant by a two-way ANOVA (*p* < 0.0001; Figure 3A). The difference in expression between the *KLHL40*^c.*152G>T^ and *KLHL40*^c.*151_*228del^ constructs was also significant (*p* < 0.0001; Figure 3A). There was no significant difference between *KLHL40*^c.*151_*228del^ and *KLHL40*^WT^ expression suggesting the *KLHL40*^c.*151_*228del^ transcript was relatively stable despite harbouring the 78 bp 3’ UTR deletion, arguing against loss of specific stabilising elements in this region.

**Figure 3.**
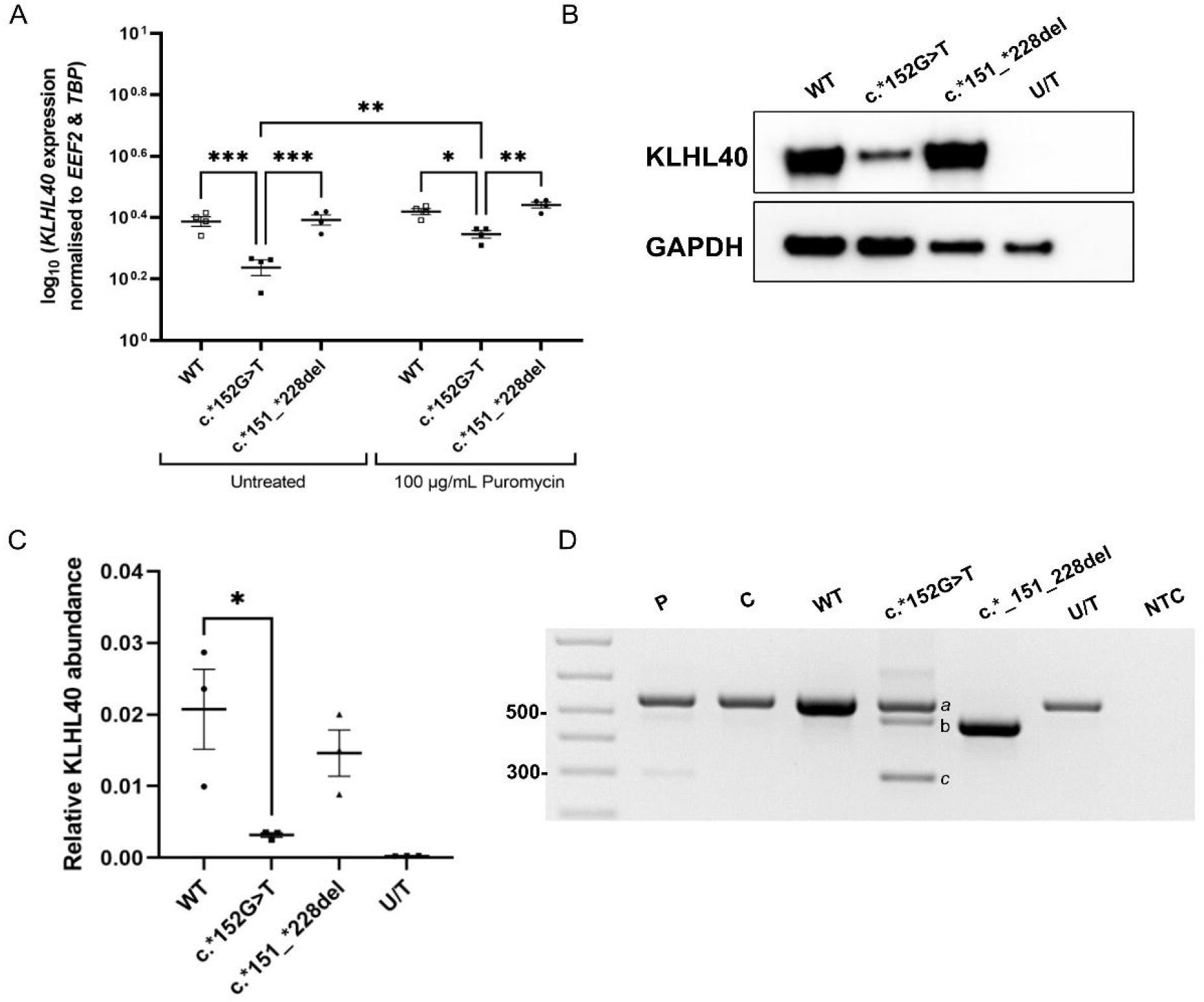
*In vitro KLHL40* expression studies. (A) log_10_ transformed qPCR data for *KLHL40* wildtype (WT), c.*152G>T, and c.*151_*228del pcDNA3.1 transcripts expressed in HEK293FT cells. Reactions performed in biological quadruplicates (*n*=4). Expression normalised to *EEF2* and *TBP*. Significance was determined by a two-way ANOVA followed by Tukey’s multiple comparison test; \**p*< 0.05, \*\**p* < 0.01, \*\*\**p* < 0.001. Error bars indicate ± SEM. (B) Representative western blot for KLHL40 from HEK293FT cells expressing wildtype (WT) and mutant (c.*152G>T, and c.*151_*228del) KLHL40 and untreated HEK293FT cells (U/T). Western blotting for GAPDH as a loading control. (C) Relative abundance of KLHL40 wildtype and mutant proteins quantified by ImageJ from KLHL40 western blots (*n*=3). Relative abundance normalised to GAPDH. Significance was determined by a one-way ANOVA followed by Tukey’s multiple comparison test; \**p* < 0.05. Error bars indicate ± SEM. (D) RT-PCR and agarose gel electrophoresis of *KLHL40* 3’ UTR products from cDNA of patient (P) and control (C) skeletal muscle, HEK293FT expressing the KLHL40 pcDNA3.1 transcripts, untreated (U/T) HEK293FT and a no template control (NTC).

To investigate whether the reduced expression of *KLHL40*^c.*152G>T^ was a consequence of nonsense mediated decay, we expressed the wildtype and mutant *KLHL40* constructs in the presence of puromycin, a known nonsense mediated decay inhibitor(16). Analyses of the log_10_-transformed expression data indicated an increase in expression of puromycin-treated *KLHL40*^c.*152G>*T*^ (2.2 ± 0.1) compared to untreated *KLHL40*^c.*152G>T^ (1.7 ± 0.1; Figure 3A). The increase in expression of *KLHL40*^c.*152G>T^ was statistically significant by a two-way ANOVA (*p* = 0.005; Figure 3A) suggesting that the splice-altering transcript generated from the c.*152G>T variant undergoes nonsense mediated decay.

Western blotting analyses indicated significantly lower abundance of KLHL40^c.*152G>T^ corresponding to ∼15% of the abundance of KLHL40^WT^ (Figure 3B,C). The difference was statistically significant by a one-way ANOVA (*p* = 0.036) and further supports an association of the c.*152G>T variant with reduced KLHL40 abundance (Figure 3C). While each experiment indicated lower abundance of KLHL40^c.*152G>T^ compared to KLHL40^c.*151_*228del^ (Figure 3B,C), the difference was not statistically significant by a one-way ANOVA.

RT-PCR analysis of the *KLHL40* 3’ UTR from patient skeletal muscle cDNA and *KLHL40*^c.*152G>T^ cDNA revealed three distinct products (Figure 3D). The largest and most prominent product (*a*) was of comparable size to *KLHL40*^WT^ (551 bp; Figure 3D) and corresponds to the un-spliced transcript with the single nucleotide variant (Supplementary Fig 1). Products *b* and *c* were relatively less prominent in the patient and *KLHL40*^c.*152G>T^ samples and were undetected in control skeletal muscle cDNA and *KLHL40*^WT^ cDNA (Figure 3D). Product *b* was of comparable size to *KLHL40*^c.*151_*228del^ (473 bp; Figure 3D) and likely corresponds to a spliced transcript containing the 78 bp deletion (Supplementary Fig 1). The smallest product (*c*; ∼300 bp) appears to correspond to the cryptic transcript predicted by SpliceAI and Introme (Figure 3D). Sequencing alignments confirmed this product contained a 255 bp deletion spanning positions c.*151 to c.*405 (Supplementary Fig 1). This suggests that in addition to the 78 bp deletion detected by RNA-seq (c.*151_*228del), the primary c.*152G>T variant also induces splicing of a 255 bp cryptic intron which results in a second deletion (c.*151_*405del) in the 3’ UTR of *KLHL40* in the patient.

### Analysis of 3’ UTR variants in ClinVar

To estimate the occurrence of 3’ UTR splicing as a mechanism of rare disease, we interrogated the ClinVar(17) database (n=1,113,674 variants, GRCh38 VCF dated 22/01/2022) for non-benign 3’ UTR variants with permissive SpliceAI scores (≥ 0.1) similar to that of *KLHL40* c.*152G>T. Our analyses indicated that 400 of 33,237 (ie ∼1.2%) 3’ UTR variants in canonical transcripts have SpliceAI scores ≥ 0.1, as presented in Supplementary table 1. Some of these variants have been reported in the literature (Table 1) and support that 3’ UTR splicing may be an underappreciated mechanism of Mendelian disease.

**Table 1.**
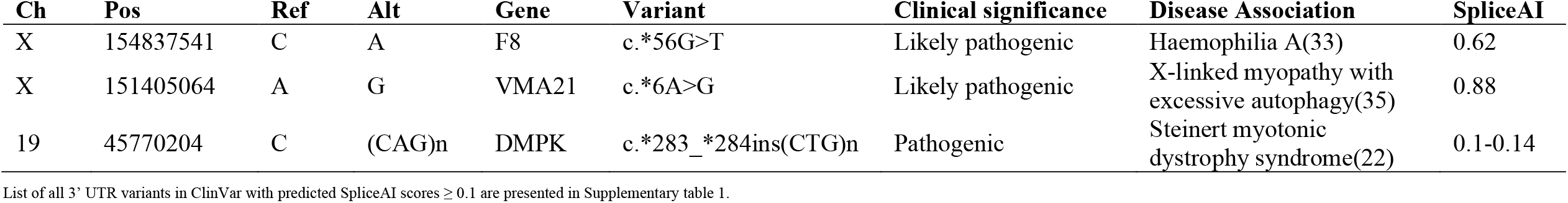
Highlighted ClinVar variants that may be associated with 3’ UTR splicing-provoked nonsense mediated decay.

| Ch | Pos | Ref | Alt | Gene | Variant | Clinical significance | Disease Association | SpliceAI |
| --- | --- | --- | --- | --- | --- | --- | --- | --- |
| X | 154837541 | C | A | F8 | c.*56G>T | Likely pathogenic | Haemophilia A(33) | 0.62 |
| X | 151405064 | A | G | VMA21 | c.*6A>G | Likely pathogenic | X-linked myopathy with excessive autophagy(35) | 0.88 |
| 19 | 45770204 | C | (CAG)n | DMPK | c.*283_*284ins(CTG)n | Pathogenic | Steinert myotonic dystrophy syndrome(22) | 0.1-0.14 |

List of all 3' UTR variants in ClinVar with predicted SpliceAI scores $\geq 0.1$ are presented in Supplementary table 1.

## Discussion

We describe a patient with NEM8 with two novel variants in *KLHL40*, a 10.9 kb deletion spanning exons 2-6 and a single nucleotide 3’ UTR variant (c.*152G>T). Together, these biallelic variants fit with the recessive inheritance pattern of NEM8(6). Notably, the patient’s clinical phenotype is milder than most NEM8 cases described in the literature(6, 7, 18). He represents one of very few cases who survived into adolescence(6) and remained ambulant, albeit with walking difficulties until adolescence. The findings in his skeletal muscle biopsy, including nemaline bodies supported his NEM8 diagnosis. It is noteworthy that intranuclear rods were also observed on biopsy, given this feature has not been previously reported in NEM8 cases. The patient’s skeletal muscle revealed significantly low levels of KLHL40 compared to controls (Figure 2B), thus supporting *KLHL40* as the underlying genetic cause of his disease.

The exon 2-6 deletion is expected to generate a non-functional transcript and therefore no functional KLHL40 protein will be produced from that allele. Given the biallelic nature of *KLHL40*-related disease(6), we believe the c.*152G>T variant in *KLHL40* is the second underlying cause of nemaline myopathy in the patient based on the following evidence. This variant was detected *in trans* with the pathogenic deletion and was predicted by SpliceAI and Introme to create a cryptic donor splice site in the 3’ UTR. Splicing within the 3’ UTR has been suggested to reduce transcript stability and abundance(19, 20). Consistently, our data from RNA-seq and western blotting of patient skeletal muscle respectively indicated a significant reduction in *KLHL40* mRNA and protein compared to controls. These findings were reproduced by our *in vitro* studies of *KLHL40*^c.*152G>T^ and suggest that the c.*152G>T variant is likely pathogenic. Of note, the low frequency of the c.*152G>T variant in the healthy population suggests that this variant could become a recurrent cause of NEM8 and should be considered in patients presenting with nemaline myopathy.

Interestingly, the c.*152G>T variant appeared to induce multiple splicing events in the 3’ UTR. Splicing of the 78 bp cryptic intron generates a deletion; c.*151_*228del that was detected in patient RNA-seq data (Figure 2F). The 78 bp deletion was also apparent in RT-PCR studies of both patient skeletal muscle cDNA and *KLHL40*^c.*152G>T^ expression construct cDNA and was undetected in controls (Figure 3D). Although the 255 bp deletion predicted by SpliceAI and Introme was undetected by RNA-seq, it was apparent by RT-PCR of both patient skeletal muscle cDNA and *KLHL40*^c.*152G>T^ construct cDNA but was undetected in controls (Figure 3D). We predict that the 255 bp deletion may prevent maturation of the resulting transcript, which may explain why it was undetected by RNA-seq.

We initially postulated that the 3’ UTR deletions would reduce *KLHL40* stability by removing recognition sites for regulatory elements such as polyadenylation proteins(21, 22), or by introducing illegitimate microRNA sites(23, 24). However, there were no remarkable annotations for the deleted regions in 3’ UTR databases. Moreover, our *in vitro* expression analyses suggested that the 78 bp 3’ UTR deletion itself does not significantly affect expression of *KLHL40* (Figure 3A-C). While mRNA and protein abundance following expression of *KLHL40*^c.*152G>T^ were significantly lower than wildtype *KLHL40*, mRNA and protein abundance following expression of *KLHL40*^c.*151_*228del^ appeared comparable to the wildtype despite containing the 78 bp deletion (Figure 3A-C). This suggested that the c.*152G>T variant was the driving cause of reduced transcript levels and protein abundance. Attenuation of nonsense mediated decay by puromycin treatment led to significantly increased expression of *KLHL40*^c.*152G>T^ (Figure 3A). Taken together, we propose that the pathomechanism of c.*152G>T may be attributed to a rare form of nonsense mediated decay provoked by 3’ UTR intron splicing(19, 25). Our data suggests that introns in UTRs are significant modulators of gene expression warranting further recognition(19).

Nonsense mediated decay is implicated in various genetic disorders(26). In most reports, this RNA surveillance mechanism is triggered by variants that result in protein pretermination, including frameshift, nonsense, and splice variants. A rarer mechanism of nonsense mediated decay involves splicing in the 3’ UTR has been proposed in domain specific reviews e.g. (19). It has been suggested that intronic insertions into most human 3’ UTRs at a distance >50–55 bp downstream of the termination codon will stimulate nonsense mediated decay(20). This was functionally demonstrated by introduction of a spliceable intron 62 bp into the 3’ UTR of wild-type β-Globin mRNA which reduced protein abundance to 24% of the wildtype(27). Notably, while this mechanism has been demonstrated by *in vitro* studies(27-29), to our knowledge, nonsense mediated decay provoked by 3’ UTR splicing has not been reported as a mechanism of Mendelian disease to date.

3’ UTR-splice activated nonsense mediated decay may explain the patient’s relatively milder disease presentation. Given the c.*152G>T cryptic donor splice site is weakly spliced, a proportion of transcripts are expected to evade aberrant splicing and proceed to translation of functional KLHL40 in the patient. Indeed, a proportion of the canonical *KLHL40* transcript was detected by RNA-seq of patient skeletal muscle (Figure 2F) as well as in our *in vitro* studies of *KLHL40*^c.*152G>T^ (Figure 3).

Mechanistically, 3’ UTR splice-activated nonsense mediated decay involves deposition of RNA binding complexes, such as the human UPF protein complex and the exon junction complex at the 3’ UTR(25, 28). Following translation termination, interactions between the 3’ UTR-bound protein complexes and the terminating ribosome signals activate nonsense mediated decay(19, 26, 30). We postulate that the cryptic introns introduced by the c.*152G>T variant may influence recruitment of such complexes to the 3’ UTR and thus trigger nonsense mediated decay(25).

Despite the crucial roles of 3’ UTRs in mRNA metabolism and surveillance(19, 28, 31), variants identified in 3’ UTRs are often overlooked during disease gene screening(23). This may be due to limited guidelines available for the clinical interpretation and functional validation of 3’ UTR variants(5). While there are some reports of pathogenic 3’ UTR variants mediating aberrant splicing(23, 32, 33), the mechanism of nonsense mediated decay provoked by 3’ UTR splicing appears to be underrecognised. Notably, a 3’ UTR variant in *F8* (c.*56G>T) has been similarly reported to create a cryptic donor splice site that results in a 159 bp deletion in the 3’ UTR (Table 1)(33). This deletion was associated with significantly reduced *F8* mRNA levels and reported to cause a milder form of haemophilia A(33). It is tempting to suggest that the underlying mechanism of this *F8* c.*56G>T variant also involves 3’ UTR splice-activated nonsense-mediated decay, particularly given it conforms with the >50–55 bp of termination codon rule(20). Additionally, 3’ UTR (CTG)n expansions in the DM1 Protein Kinase gene (*DMPK*) are a cause of *DMPK* deficiency in patients with myotonic dystrophy (OMIM# 160900). There are several speculations as to how the expansions reduce *DMPK* mRNA and protein abundance, one of which involves aberrant splicing(22, 34). Overall, such reports indicate that aberrant 3’ UTR-splicing may underlie genetically unresolved rare diseases.

Our analysis of the ClinVar database (n=1,113,674 variants. GRCh38 VCF dated 22/01/2022) suggested a small percentage (∼1.2%) of 3’ UTR variants could induce splicing (Supplementary table 1). We predict that the prevalence of this mechanism may be underrepresented given that 3’ UTRs are not usually interrogated during variant curation(5) and thus 3’ UTR variants are less likely to be submitted to ClinVar. Of the 3’ UTR variants that have been submitted, few appear to have been functionally investigated. Moreover, the mechanism for some of the 3’ UTR variants that have been investigated remains unknown(35, 36). This includes a c.*6A>G variant in *VMA21* associated with X-linked myopathy with excessive autophagy (OMIM# 310440, Table 1). This variant was reported twice in ClinVar (VCV000208803.4) and in 20 patients from three families(35). While Ramachandran *et al*.(35) functionally showed that the c.*6A>G variant was associated with reduced *VMA21* transcript and protein abundance, the underlying mechanism remains unconfirmed. Our SpliceAI analyses suggest that the c.*6A>G substitution may lead to the creation of a new donor splice site at position c.*1 with a high donor gain prediction score of 0.88 (Δ ≥ 0.8)(13). Although Ramachandran *et al*.(35) did not detect aberrant splicing at this position by RT-PCR of patient lymphoblasts, it would be interesting to investigate the *VMA21* 3’ UTR by an alternative method such as RNA-seq to determine whether the c.*6A>G variant is another example of pathogenic 3’ UTR splicing(37). Overall, it is apparent that 3’ UTR variants warrant further recognition during variant curation(19, 24). We encourage submission of 3’ UTR variants to disease gene databases to facilitate improved curation and classification of 3’ UTR variants in Mendelian disease.

## Conclusions

This study identified a likely pathogenic splice variant in the 3’ UTR of *KLHL40* (c.*152G>T) in a patient with a milder presentation of NEM8. This finding expands the genotypic and phenotypic spectrum of NEM8. Using next generation sequencing, RNA-seq, and *in vitro* studies, we provide the first evidence of a pathogenic variant destabilising the *KLHL40* mRNA by 3’ UTR splice-activated nonsense mediated decay. Aberrant 3’ UTR splicing may be an underrecognised mechanism of Mendelian disease. Our study highlights the utility of RNA-seq in 1) detecting expression outliers and 2) identifying cryptic splice variants in non-coding genomic regions that may evade detection and/or curation in data generated from targeted gene panel and whole exome sequencing(37, 38). Inclusion of RNA-seq into the diagnostic toolkit for rare disorders can thus enable identification of pathogenic variants that may contribute to the number of cases remaining undiagnosed following genetic screening(37). Overall, this study highlights the necessity of looking beyond the coding regions when investigating the molecular basis of rare disease(24, 33).

## Patient and methods

This project was approved by the University of Western Australia Human Research Ethics Committee (RA/4/20/1008) and the Curtin University Human Research Ethics Office (HRE2019-0566). Written informed consent was provided by the family to have the results of this research published.

### Patient details

The patient was diagnosed with nemaline myopathy on the basis of clinical findings and histopathological findings on skeletal muscle biopsy. Clinical assessments, MRI, muscle biopsy analysis and western blotting of patient muscle biopsy were performed at the Biomedicine Institute of Sevilla and the Department of Neurology at Virgen del Rocío University Hospital in Sevilla, Spain.

### Methods for histology and electron microscopy

Muscle biopsy was obtained from the quadriceps muscle, immediately frozen by standard methods, and processed following the standard procedures(39). Routine histochemical techniques were performed on 7 μM transverse sections of frozen muscle, including hematoxylin and eosin, modified Gomori trichrome, oil red O, periodic acid-Schiff, nicotinamide adenine dinucleotide-tetrazolium reductase (NADH-TR), succinate dehydrogenase (SDH), cytochrome C oxidase (COX), and adenosine triphosphatase (ATPase) pH 9.4, 4.6, and 4.3.

Ultrastructural studies were performed following standard methods(39). A small fragment of the muscle was fixed in 2.5% w/v glutaraldehyde solution, postfixed in 1% w/v osmium tetroxide, and embedded in epoxy resin. Semithin sections were stained with 1% toluidine blue. Ultrathin sections were mounted on copper grids and examined with a Zeiss Libra 120 transmission electron microscope (Carl Zeiss NTS GmbH, Oberkochen, Germany).

### Western blotting

Frozen muscle samples were homogenized in RIPA buffer (20 mM Tris HCl pH 7.4, 150 mM NaCl, 1 mM EDTA, 1% v/v IGEPAL, 0.1% w/v SDS) containing protease inhibitor cocktail (Roche). The lysates were centrifuged at 15,781 g at 4°C for 20 min. The supernatant was collected. The protein lysates were separated on 10% w/v SDS-PAGE gels and transferred onto PDVF membranes (Millipore). Western blot analysis of equal-protein loading was performed with the following primary antibodies: rabbit polyclonal anti-KLHL40 (Sigma-Aldrich; HPA024463; [1:500]) and rabbit polyclonal anti-GAPDH (Sigma-Aldrich; [1:2,000]). Immunoreactivity was detected with secondary antibodies conjugated to horseradish peroxidase (1:5,000; Jackson Immuno Research) and developed with SuperSignal West Femto (Thermo Fisher Scientific) using an ImageQuant LAS 4000 MiniGold System (GE Healthcare Life Sciences).

### Genetic analyses

#### Neuromuscular disease gene panel

DNA from the proband was sequenced on the neuromuscular disease gene panel at PathWest using Illumina sequencing chemistry as previously described(4, 40). Base calling, mapping, and variant calling were performed as previously described(4, 40).

#### Whole exome sequencing

Illumina whole exome sequencing (WES) was performed at the Broad Institute of Harvard and MIT as previously described(4). Exome data were analysed in seqr(12) (https://seqr.broadinstitute.org).

#### Variant interpretation and confirmation

Variants were analysed and interpreted following standard bioinformatic approaches(41, 42). Candidate single nucleotide variants were validated by bidirectional Sanger sequencing (Australian Genome Research Facility (AGRF), Perth)(40). Sanger chromatograms were aligned to a reference sequence in benchling (benchling.com) using the MAFFT algorithm.

#### In silico variant prediction analysis

Introme(14) (https://github.com/CCICB/introme; manuscript in preparation) and SpliceAI(13) (https://spliceailookup.broadinstitute.org/) were used to predict the effect of variants on splicing.

#### Analysis of 3’ UTR variants in ClinVar

Variants were downloaded from ClinVar(17) (GRCh38, 22/01/2022; https://ftp.ncbi.nlm.nih.gov/pub/clinvar/vcf_GRCh38/archive_2.0/2022/clinvar_20220122.vcf.gz) in VCF format (n=1,113,674 variants). BCFtools(43) query was used to extract Class 3-5 variants (VUS, likely pathogenic and pathogenic; n=623,975 variants). BCFtools view was used to extract variants with a Molecular Consequence of ‘3_prime_UTR_VARIANT’ (n=43,775 variants). These variants were run on SpliceAI(13). Eleven variants had reference alleles that were deemed too long to annotate by SpliceAI and variants with no annotation were removed, leaving 38,515 variants. Next, the VCF was annotated using VEP(44) to determine the canonical transcript for each variant. Variants in the 3’ UTR of the respective canonical transcript were retained, leaving 33,237 variants in the 3’ UTR of the canonical transcript. We chose a SpliceAI threshold of 0.10 to keep variants, based on the *KLHL40* c.*152G>T score (0.17) and the optimal threshold (0.11) reported by Riepe *et al*.(45) for their *MYBPC3* dataset, in contrast to the SpliceAI suggested threshold of 0.20(13). These data are presented in Supplementary Table 1.

#### RNA sequencing

RNA extraction from patient skeletal muscle and RNA sequencing were performed at the Department of Diagnostic Genomics (PathWest, Perth) as previously described(46). RNA-seq data were visualised with the Integrative Genomics Viewer (IGV)(47). Data were also assembled and analysed using MINTIE(48), a reference-free RNA-seq analysis pipeline that maps reads *de novo* to identify novel structural and splice variants(15). Muscle RNA-seq FASTQ data from five additional non-nemaline myopathy patients were used as controls for MINTIE.

### *In vitro* expression analyses

#### Expression constructs

Mammalian pcDNA3.1 expression constructs (pcDNA3.1) were synthesised by Genscript. These constructs contained the KLHL40 coding sequence (ENST00000287777) with the wild-type or mutant 3’ UTR encoding either of two variants: c.*152G>T or c.*151_*228del.

#### Cell culture

Human embryonic kidney cell lines (HEK293FT) were maintained in Dulbecco’s Modified Eagle Medium (DMEM; Gibco) supplemented with 10% fetal bovine serum (FBS) and 1% Penicillin/Streptomycin (Gibco). At 50-60% confluency, expression constructs were transfected following the Viafect kit protocol (Promega) at a 4:1 ratio (Viafect transfection reagent: DNA). Cultures were maintained at 37°C with 5% CO_2_. A pmaxGFP vector was transfected alongside to estimate transfection efficiency. Cells were harvested at 72 h post-transfection and pellets snap frozen and stored at -80°C until required for RNA and protein extraction.

##### Puromycin assays

HEK293FT cells were maintained in DMEM + 10% FBS and transfected as above. At 21 h post-transfection, cells were incubated with media containing either 100 µg/mL puromycin dihydrochloride (Thermo Fisher [# A1113803]) or no puromycin for 3 h at 37°C with 5% CO_2_ before harvesting at 24 h post-transfection.

#### RNA extraction and quantitative PCR

Total RNA was extracted from HEK293FT pellets using the RNeasy mini kit protocol (Qiagen). Complementary DNA (cDNA) was synthesised from 1 µg RNA using the SuperScript III First-Strand Synthesis System (Thermo Fisher Scientific, Waltham, MA). To investigate relative transcript expression levels of wildtype *KLHL40, KLHL40*^c.*152G>T^ and *KLHL40*^c.*151_*228del^, qPCR was performed as previously described(49). Reactions were performed in 10 µL volumes containing a 2X Rotor-Gene SYBR Green PCR master mix (Qiagen; 204076), 1 µL diluted cDNA and 0.8 μM of the following forward and reverse primers: KLHL40_FD5_6 (5’ GGAGGTATAACGAGGAGGAGAA-3’, bridges the junction of exon 5 and exon 6), and KLHL40_RV6 (5’-CTGAGCTGGTCACATCTTAGTC-3’). Reactions were performed in technical duplicates for each biological replicate (*n*=4) and Ct values were averaged. Expression was normalised using the delta Ct method(50), comparing to the geometric mean of two reference genes (*TBP* and *EEF2*)(49). Data were log_10_ transformed and analysed by a two-way analysis of variance (ANOVA) followed by Tukey’s multiple comparison test to measure statistical significance using GraphPad Prism 9.0.1 (La Jolla, USA).

#### Reverse transcriptase PCR

RT-PCR was performed to confirm the occurrence of mis-splicing events in the 3’ UTR region. Reactions were performed in 25 µL volumes containing a 1X GoTaq G2 master mix (Promega, Madison, WI [#M7832]), 0.4 µM forward primer (KLHL40_UTR_F; 5’-CCAGCTCAGGCAGACTGAAC-3’), 0.4 µM reverse primer (KLHL40_UTR_R; 5-GACACCAGATGGAGAGCAGAG-3’) and 7.5 ng cDNA. Touchdown PCR cycling conditions were: 5 min at 98°C; 15 cycles of 15 s at 98°C, 10 s at 65°C-57.5°C (−0.5°C per cycle), and 15 s at 72°C; 30 cycles of 15 s at 98°C, 10 s at 57°C, and 15 s at 72°C; and a final cycle of 5 min at 72°C. Products were resolved on 2% w/v agarose gel in 1X TAE at 95 V for 1 h. PCR products were gel excised and purified using the QIAquick gel and PCR purification kit. DNA was quantified on a NanoDrop One spectrophotometer (Thermo Scientific) and Sanger sequenced at AGRF (Perth, Australia). Sanger chromatograms were aligned to a reference sequence in benchling (benchling.com) using the MAFFT algorithm.

#### Protein extraction and western blotting

Frozen cell pellets were suspended in lysis buffer containing 2% w/v SDS, 125 mM Tris (pH 6.8), and 1x PIC (Halt™ Protease Inhibitor Cocktail; Thermo Scientific) and homogenised by sonication. Protein concentrations were determined by a BCA protein assay (Pierce; #23227). Lysates were prepared in loading buffer containing 10% w/v SDS, 312.5 mM Tris (pH 6.8), 50% v/v glycerol, bromophenol blue saturated solution, 50 mM DTT and 1x PIC. Western blotting was performed as previously described(6). Lysates (500 ng) were separated on 4-12% w/v Bis-Tris NuPAGE Novex polyacrylamide gels and transferred to polyvinylidene difluoride transfer membranes (Thermo Fisher Scientific). After 1 h blocking with PBS + 0.1% v/v Tween-20 and 5% w/v skim milk, membranes were incubated with a Human Protein Atlas rabbit polyclonal KLHL40 (KBTBD5) antibody (Sigma-Aldrich; #HPA024463; [1:2500]) overnight at 4°C and a secondary goat anti-rabbit horseradish peroxidase antibody (Sigma; #A0545) at room temperature for 1 h. Membranes were imaged by chemiluminescent detection (Pierce ECL Plus kit; #32132) using the Invitrogen iBright FL1000 imaging system (Thermo Fisher Scientific). Membranes were also blotted with an anti-GAPDH antibody (Sigma; #G8795; [1:10,000]) as a loading control. Relative KLHL40 abundance was quantified using ImageJ(51). Statistical significance was measured using a one-way ANOVA followed by Tukey’s multiple comparisons test with a single pooled variance (*n*=3) on GraphPad Prism 9.0.1 (La Jolla, USA).

## Data Availability

All data produced in the study are available upon reasonable request.

## Acknowledgements

We would like to express our gratitude to the patient and family for their involvement in this study. We thank Associate Professor David Groth and Dr. Danielle Dye for their inputs into this study. LD is supported by an Australian Government Research Training Program (RTP) Scholarship. GR (Investigator Grant, APP2007769) and NGL (Fellowship APP1117510) are supported by the Australian NHMRC. This work is funded by NHMRC Ideas Grant (APP2002640). This work was supported by resources provided by the Pawsey Supercomputing Centre with funding from the Australian Government and the Government of Western Australia.

## Legends to Figures

**Supplementary Fig 1. Sanger sequencing of RT-PCR products from *in vitro* expression of KLHL40**^**WT**^ **and KLHL40**^**c.*152G>T**^. (A) Close up view and (D) expanded view of sequence chromatograms. Corresponding gel products are shown in Figure 3D.

## Abbreviations

IGV: Integrative Genomics Viewer
NEM8: nemaline myopathy 8
RT-PCR: reverse transcriptase polymerase chain reaction

## References

1 Nowak, K.J., Davis, M.R., Wallgren-Pettersson, C., Lamont, P.J. and Laing, N.G. (2012) Clinical utility gene card for: Nemaline myopathy. Eur J Hum Genet, 20, 713–713.

2 Gonorazky, H.D., Bönnemann, C.G. and Dowling, J.J. (2018) The genetics of congenital myopathies. Handb Clin Neurol, 148, 549–564.

3 Benarroch, L., Bonne, G., Rivier, F. and Hamroun, D. (2020) The 2021 version of the gene table of neuromuscular disorders (nuclear genome). Neuromuscul Disord, 30, 1008–1048.

4 Beecroft, S.J., Yau, K.S., Allcock, R.J.N., Mina, K., Gooding, R., Faiz, F., Atkinson, V.J., Wise, C., Sivadorai, P., Trajanoski, D. et al. (2020) Targeted gene panel use in 2249 neuromuscular patients: the Australasian referral center experience. Ann Clin Transl Neurol, 7, 353–362.

5 Ellingford, J.M., Ahn, J.W., Bagnall, R.D., Baralle, D., Barton, S., Campbell, C., Downes, K., Ellard, S., Duff-Farrier, C., FitzPatrick, D.R. et al. (2021) Recommendations for clinical interpretation of variants found in non-coding regions of the genome. medRxiv, in press., 2021.2012.2028.21267792.

6 Ravenscroft, G., Miyatake, S., Lehtokari, V.-L., Todd Emily J., Vornanen, P., Yau Kyle S., Hayashi Yukiko K., Miyake, N., Tsurusaki, Y., Doi, H. et al. (2013) Mutations in KLHL40 Are a Frequent Cause of Severe Autosomal-Recessive Nemaline Myopathy. Am J Hum Genet, 93, 6–18.

7 Chen, T.H., Tian, X., Kuo, P.L., Pan, H.P., Wong, L.C. and Jong, Y.J. (2016) Identification of KLHL40 mutations by targeted next-generation sequencing facilitated a prenatal diagnosis in a family with three consecutive affected fetuses with fetal akinesia deformation sequence. Prenat Diagn, 36, 1135–1138.

8 Seferian, A.M., Malfatti, E., Bosson, C., Pelletier, L., Taytard, J., Forin, V., Gidaro, T., Gargaun, E., Carlier, P., Fauré, J. et al. (2016) Mild clinical presentation in KLHL40-related nemaline myopathy (NEM 8). Neuromuscul Disord, 26, 712–716.

9 Natera-de Benito, D., Nascimento, A., Abicht, A., Ortez, C., Jou, C., Müller, J.S., Evangelista, T., Töpf, A., Thompson, R., Jimenez-Mallebrera, C. et al. (2016) KLHL40-related nemaline myopathy with a sustained, positive response to treatment with acetylcholinesterase inhibitors. J Neurol, 263, 517–523.

10 Garg, A., O’Rourke, J., Long, C., Doering, J., Ravenscroft, G., Bezprozvannaya, S., Nelson, B.R., Beetz, N., Li, L., Chen, S. et al. (2014) KLHL40 deficiency destabilizes thin filament proteins and promotes nemaline myopathy. J Clin Invest, 124, 3529–3539.

11 Gong, W., Gohla, R.M., Bowlin, K.M., Koyano-Nakagawa, N., Garry, D.J. and Shi, X. (2015) Kelch Repeat and BTB Domain Containing Protein 5 (Kbtbd5) Regulates Skeletal Muscle Myogenesis through the E2F1-DP1 Complex. J Biol Chem, 290, 15350–15361.

12 Pais, L., Snow, H., Weisburd, B., Zhang, S., Baxter, S., DiTroia, S., O’Heir, E., England, E., Chao, K., Lemire, G. et al. (2021) seqr: a web-based analysis and collaboration tool for rare disease genomics. medRxiv, in press., 2021.2010.2027.21265326.

13 Jaganathan, K., Kyriazopoulou Panagiotopoulou, S., McRae, J.F., Darbandi, S.F., Knowles, D., Li, Y.I., Kosmicki, J.A., Arbelaez, J., Cui, W., Schwartz, G.B. et al. (2019) Predicting Splicing from Primary Sequence with Deep Learning. Cell, 176, 535-548.e524.

14 Sullivan, P., Mayoh, C., Wong-Erasmus, M., Gayevskiy, V., Beecroft, S., Pinese, M., Oates, E. and Cowley, M. (2020) P.334 Introme identifies non-canonical splice-altering variants in neuromuscular patients resulting in multiple new genetic diagnoses. Neuromuscul Disord, 30, S144.

15 Cmero, M., Schmidt, B., Majewski, I.J., Ekert, P.G., Oshlack, A. and Davidson, N.M. (2021) MINTIE: identifying novel structural and splice variants in transcriptomes using RNA-seq data. Genome Biol, 22, 296.

16 Gaildrat, P., Killian, A., Martins, A., Tournier, I., Frebourg, T. and Tosi, M. (2010) Use of splicing reporter minigene assay to evaluate the effect on splicing of unclassified genetic variants. Methods Mol Biol, 653, 249–257.

17 Landrum, M.J., Lee, J.M., Benson, M., Brown, G.R., Chao, C., Chitipiralla, S., Gu, B., Hart, J., Hoffman, D., Jang, W. et al. (2018) ClinVar: improving access to variant interpretations and supporting evidence. Nucleic Acids Res, 46, D1062–d1067.

18 Todd, E.J., Yau, K.S., Ong, R., Slee, J., McGillivray, G., Barnett, C.P., Haliloglu, G., Talim, B., Akcoren, Z., Kariminejad, A. et al. (2015) Next generation sequencing in a large cohort of patients presenting with neuromuscular disease before or at birth. Orphanet J Rare Dis, 10, 148–148.

19 Bicknell, A.A., Cenik, C., Chua, H.N., Roth, F.P. and Moore, M.J. (2012) Introns in UTRs: Why we should stop ignoring them. BioEssays, 34, 1025–1034.

20 Nagy, E. and Maquat, L.E. (1998) A rule for termination-codon position within intron-containing genes: when nonsense affects RNA abundance. Trends Biochem Sci, 23, 198–199.

21 Nourse, J., Spada, S. and Danckwardt, S. (2020) Emerging Roles of RNA 3’-end Cleavage and Polyadenylation in Pathogenesis, Diagnosis and Therapy of Human Disorders. Biomolecules, 10.

22 Frisch, R., Singleton, K.R., Moses, P.A., Gonzalez, I.L., Carango, P., Marks, H.G. and Funanage, V.L. (2001) Effect of Triplet Repeat Expansion on Chromatin Structure and Expression of DMPK and Neighboring Genes, SIX5 and DMWD, in Myotonic Dystrophy. Mol Genet Metab, 74, 281–291.

23 Liaqat, K., Chiu, I., Lee, K., Chakchouk, I., Andrade-Elizondo, P.B., Santos-Cortez, R.L.P., Hussain, S., Nawaz, S., Ansar, M., Khan, M.N. et al. (2018) Novel missense and 3’-UTR splice site variants in LHFPL5 cause autosomal recessive nonsyndromic hearing impairment. J Hum Genet, 63, 1099–1107.

24 Dusl, M., Senderek, J., Müller, J.S., Vogel, J.G., Pertl, A., Stucka, R., Lochmüller, H., David, R. and Abicht, A. (2015) A 3’-UTR mutation creates a microRNA target site in the GFPT1 gene of patients with congenital myasthenic syndrome. Hum Mol Genet, 24, 3418–3426.

25 Giorgi, C., Yeo, G.W., Stone, M.E., Katz, D.B., Burge, C., Turrigiano, G. and Moore, M.J. (2007) The EJC factor eIF4AIII modulates synaptic strength and neuronal protein expression. Cell, 130, 179–191.

26 Kurosaki, T. and Maquat, L.E. (2016) Nonsense-mediated mRNA decay in humans at a glance. J Cell Sci, 129, 461–467.

27 Thermann, R., Neu-Yilik, G., Deters, A., Frede, U., Wehr, K., Hagemeier, C., Hentze, M.W. and Kulozik, A.E. (1998) Binary specification of nonsense codons by splicing and cytoplasmic translation. EMBO J, 17, 3484–3494.

28 Lykke-Andersen, J., Shu, M.D. and Steitz, J.A. (2000) Human Upf proteins target an mRNA for nonsense-mediated decay when bound downstream of a termination codon. Cell, 103, 1121–1131.

29 Carter, M.S., Li, S. and Wilkinson, M.F. (1996) A splicing-dependent regulatory mechanism that detects translation signals. EMBO J, 15, 5965–5975.

30 Le Hir, H., Gatfield, D., Izaurralde, E. and Moore, M.J. (2001) The exon-exon junction complex provides a binding platform for factors involved in mRNA export and nonsense-mediated mRNA decay. EMBO J, 20, 4987–4997.

31 Preussner, M., Gao, Q., Morrison, E., Herdt, O., Finkernagel, F., Schumann, M., Krause, E., Freund, C., Chen, W. and Heyd, F. (2020) Splicing-accessible coding 3′UTRs control protein stability and interaction networks. Genome Biol, 21, 186.

32 Jin, C., Jiang, J., Wang, W. and Yao, K. (2010) Identification of a MIP mutation that activates a cryptic acceptor splice site in the 3’ untranslated region. Mol Vis, 16, 2253–2258.

33 Pezeshkpoor, B., Berkemeier, A.C., Czogalla, K.J., Oldenburg, J. and El-Maarri, O. (2016) Evidence of pathogenicity of a mutation in 3′ untranslated region causing mild haemophilia A. Haemophilia, 22, 598–603.

34 Carango, P., Noble, J.E., Marks, H.G. and Funanage, V.L. (1993) Absence of myotonic dystrophy protein kinase (DMPK) mRNA as a result of a triplet repeat expansion in myotonic dystrophy. Genomics, 18, 340–348.

35 Ramachandran, N., Munteanu, I., Wang, P., Ruggieri, A., Rilstone, J.J., Israelian, N., Naranian, T., Paroutis, P., Guo, R., Ren, Z.P. et al. (2013) VMA21 deficiency prevents vacuolar ATPase assembly and causes autophagic vacuolar myopathy. Acta Neuropathol, 125, 439–457.

36 Ruggieri, A., Ramachandran, N., Wang, P., Haan, E., Kneebone, C., Manavis, J., Morandi, L., Moroni, I., Blumbergs, P., Mora, M. et al. (2015) Non-coding VMA21 deletions cause X-linked myopathy with excessive autophagy. Neuromuscul Disord, 25, 207–211.

37 Cummings, B.B., Marshall, J.L., Tukiainen, T., Lek, M., Donkervoort, S., Foley, A.R., Bolduc, V., Waddell, L.B., Sandaradura, S.A., O’Grady, G.L. et al. (2017) Improving genetic diagnosis in Mendelian disease with transcriptome sequencing. Sci Transl Med, 9, 1–11.

38 Gonorazky, H.D., Naumenko, S., Ramani, A.K., Nelakuditi, V., Mashouri, P., Wang, P., Kao, D., Ohri, K., Viththiyapaskaran, S., Tarnopolsky, M.A. et al. (2019) Expanding the Boundaries of RNA Sequencing as a Diagnostic Tool for Rare Mendelian Disease. Am J Hum Genet, 104, 466–483.

39 Dubowitz, V., Sewry, C.A. and Oldfors, A. (2020) Muscle biopsy A Practical Approach. Elsevier, London.

40 Ravenscroft, G., Clayton, J.S., Faiz, F., Sivadorai, P., Milnes, D., Cincotta, R., Moon, P., Kamien, B., Edwards, M., Delatycki, M. et al. (2021) Neurogenetic fetal akinesia and arthrogryposis: genetics, expanding genotype-phenotypes and functional genomics. J Med Genet, 58, 609–618.

41 Richards, S., Aziz, N., Bale, S., Bick, D., Das, S., Gastier-Foster, J., Grody, W.W., Hegde, M., Lyon, E., Spector, E. et al. (2015) Standards and guidelines for the interpretation of sequence variants: a joint consensus recommendation of the American College of Medical Genetics and Genomics and the Association for Molecular Pathology. Genet Med, 17, 405–423.

42 Dashti, M.J.S. and Gamieldien, J. (2017) A practical guide to filtering and prioritizing genetic variants. BioTechniques, 62, 18–30.

43 Danecek, P., Bonfield, J.K., Liddle, J., Marshall, J., Ohan, V., Pollard, M.O., Whitwham, A., Keane, T., McCarthy, S.A., Davies, R.M. et al. (2021) Twelve years of SAMtools and BCFtools. Gigascience, 10.

44 McLaren, W., Gil, L., Hunt, S.E., Riat, H.S., Ritchie, G.R., Thormann, A., Flicek, P. and Cunningham, F. (2016) The Ensembl Variant Effect Predictor. Genome Biol, 17, 122.

45 Riepe, T.V., Khan, M., Roosing, S., Cremers, F.P.M. and t Hoen, P.A.C. (2021) Benchmarking deep learning splice prediction tools using functional splice assays. Hum Mutat, 42, 799–810.

46 Bryen, S.J., Ewans, L.J., Pinner, J., MacLennan, S.C., Donkervoort, S., Castro, D., Töpf, A., O’Grady, G., Cummings, B., Chao, K.R. et al. (2020) Recurrent TTN metatranscript-only c.39974-11T>G splice variant associated with autosomal recessive arthrogryposis multiplex congenita and myopathy. Hum Mutat, 41, 403–411.

47 Thorvaldsdóttir, H., Robinson, J.T. and Mesirov, J.P. (2013) Integrative Genomics Viewer (IGV): high-performance genomics data visualization and exploration. Brief Bioinform, 14, 178–192.

48 Cmero, M., Schmidt, B., Majewski, I.J., Ekert, P.G., Oshlack, A. and Davidson, N.M. (2020) MINTIE: identifying novel structural and splice variants in transcriptomes using RNA-seq data. bioRxiv, in press., 2020.2006.2003.131532.

49 McNamara, E.L., Taylor, R.L., Clayton, J.S., Goullee, H., Dilworth, K.L., Pinós, T., Brull, A., Alexander, I.E., Lisowski, L., Ravenscroft, G. et al. (2020) Systemic AAV8-mediated delivery of a functional copy of muscle glycogen phosphorylase (Pygm) ameliorates disease in a murine model of McArdle disease. Hum Mol Genet, 29, 20–30.

50 Vandesompele, J., De Preter, K., Pattyn, F., Poppe, B., Van Roy, N., De Paepe, A. and Speleman, F. (2002) Accurate normalization of real-time quantitative RT-PCR data by geometric averaging of multiple internal control genes. Genome Biol, 3, research0034.0031.

51 Schneider, C.A., Rasband, W.S. and Eliceiri, K.W. (2012) NIH Image to ImageJ: 25 years of image analysis. Nat Methods, 9, 671–675.

