## Supplementary figures and images for "A *KLHL40* 3’ UTR splice-altering variant causes milder NEM8, an under-appreciated disease mechanism"

### Supplementary Fig 1

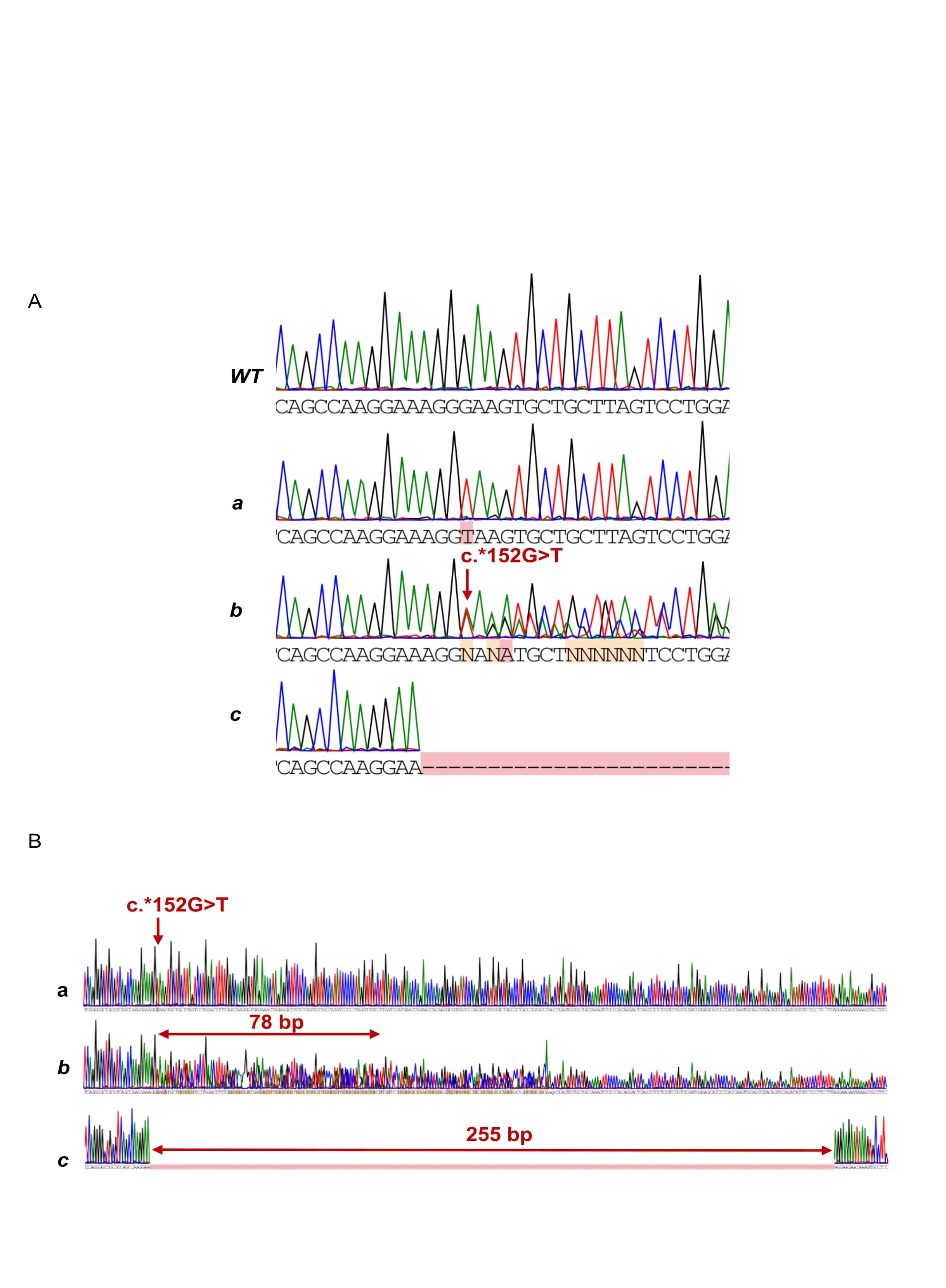
